# Epigenetic Markers of Response to Psychotherapy in Obsessive-Compulsive Disorder

**DOI:** 10.64898/2026.03.20.26348888

**Authors:** Kira D. Höffler, Anne-Kristin Stavrum, Matthew W. Halvorsen, Thorstein Olsen Eide, Kristen Hagen, Anders Lillevik Thorsen, Olga Therese Ousdal, Gerd Kvale, James J. Crowley, Jan Haavik, Kerry J. Ressler, Bjarne Hansen, Stephanie Le Hellard

## Abstract

Cognitive-behavioral therapy (CBT) is a widely used treatment for mental disorders, yet the biological mechanisms underlying its effects and the factors contributing to treatment response remain poorly understood. DNA methylation, an epigenetic mechanism shaped by both genetic and environmental influences, may provide insights into individual differences in psychotherapy outcomes. We performed an epigenome-wide association study of psychotherapy response in 889 adults with obsessive-compulsive disorder (OCD) receiving the Bergen 4-Day Treatment, a concentrated exposure-based intervention. Saliva samples were collected before treatment, immediately after treatment, and at 3-month follow-up. DNA methylation was profiled using the Illumina EPIC v2 array. We identified three classes of differentially methylated regions (DMRs): (1) ten baseline DMRs associated with subsequent treatment response, (2) 23 DMRs showing stable associations with response across time points, and (3) three DMRs exhibiting longitudinal methylation changes associated with response. These loci were annotated to genes involved in neuroplasticity, neurodevelopment, stress response, immune function, mitochondrial processes, and gene regulation. Baseline and time-stable methylation signals were frequently influenced by genetic variation, whereas longitudinal signals showed no significant local genetic effects. In addition, modest shifts in monocyte and CD4+ T-cell proportions were associated with treatment response. These findings suggest that response to exposure-based psychotherapy in OCD is associated with both trait-like and dynamic peripheral epigenetic variation. Pretreatment DNA methylation profiles may ultimately help predict treatment outcomes and support future precision psychiatry approaches.

## Introduction

Psychotherapy is a first-line treatment for many prevalent mental health disorders [1]. Yet, the biological mechanisms underlying its therapeutic effects, and the reasons why a substantial proportion of individuals fail to respond, remain poorly understood [2]. Epigenetic mechanisms, which regulate gene expression without altering the DNA sequence, are influenced by genetic, environmental, and developmental factors and therefore represent promising candidates for studying treatment response.

Most work in this field has focused on DNA methylation in peripheral tissues, involving the addition of a methyl group to DNA. Longitudinal studies have reported psychotherapy-associated changes in DNA methylation that correlate with symptom improvement [3–10], and pretreatment methylation levels have also been associated with later outcome [3,10–12], as summarized in recent reviews [13–15]. However, the current literature is limited by small sample sizes and predominantly hypothesis-driven candidate-gene approaches targeting genes such as *MAOA, NR3C1, FKBP5, OXTR,* and *SLC6A4*. Large epigenome-wide studies are therefore needed to advance understanding of treatment response biology and to identify accessible biomarkers that may support more personalized treatment.

Obsessive-compulsive disorder (OCD) is a common target for psychotherapeutic interventions. OCD substantially impairs quality of life and social, occupational, academic, and family functioning [16]. Cognitive behavioral therapy with exposure and response prevention (ERP) is the gold-standard psychotherapeutic treatment for OCD [17]. Nevertheless, a substantial proportion of patients show limited improvement [17], and predictors of treatment response remain inconsistent [18]. No clinically implemented biomarkers are currently available.

Here, we conducted a large epigenome-wide association study (EWAS) of psychotherapy response in OCD to identify DNA methylation signals associated with clinical improvement. Participants received concentrated ERP delivered as the Bergen 4-Day Treatment, an intervention with high efficacy, although up to one-quarter of patients do not achieve treatment response at follow-up [19–24]. Building on prior work, we hypothesized that pretreatment DNA methylation and longitudinal methylation change would be associated with treatment response, with some response-related signals remaining stable over time. For significant findings, we compared methylation levels with healthy controls and evaluated potential influences of genetic variation, medication use, and psychiatric comorbidities. Finally, we examined associations between estimated saliva cell-type proportions and treatment response.

## Materials and Methods

Figure 1 shows an overview of the methodological approach.

**Figure 1:**
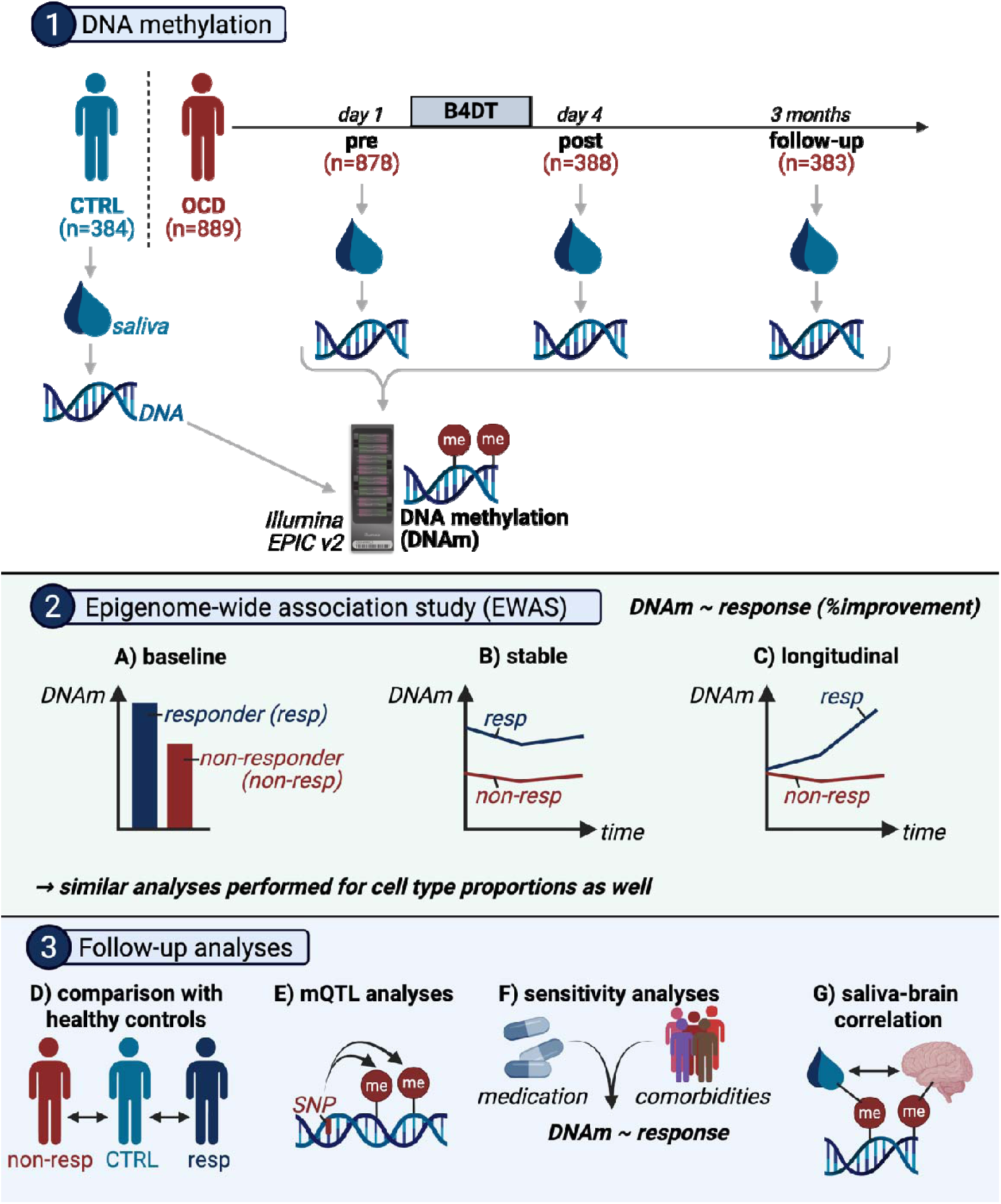
Study overview. Participant and sample numbers are reported post-quality control. Created with BioRender. B4DT: Bergen 4-Day Treatment, CTRL: control, EWAS: epigenome-wide association study, mQTL: methylation Quantitative Trait Locus, OCD: obsessive-compulsive disorder, resp: responder.

### Participants

Individuals referred for OCD treatment were recruited from eight clinics across Norway between 2016 and 2023. Eligible individuals were aged 18 years or older, fluent in Norwegian, and had a clinician-confirmed DSM-5 diagnosis of OCD [25]. Exclusion criteria included current psychotic disorder, bipolar disorder, substance use disorder, intellectual disability, severe eating disorder, or active suicidal ideation. Participants receiving psychoactive medication were encouraged to maintain a stable dosage before and during treatment. An overview of psychoactive medications is provided in **Table S1**.

Healthy control participants without self-reported OCD were drawn from a previously established Norwegian cohort [26] and matched on age (±3 years) and sex.

The study was approved by the Regional Committee for Medical and Health Research Ethics (REK Vest), and all participants provided written informed consent.

### Treatment, Clinical Assessment, and Treatment Response

Participants with OCD received the Bergen 4-Day Treatment (B4DT), a concentrated ERP-based therapy delivered in groups of three to six patients over four days with a 1:1 therapist-patient ratio. Treatment procedures have been described previously [27].

OCD symptom severity was assessed using the Yale-Brown Obsessive-Compulsive Scale (Y-BOCS) [28] at baseline, ten days after treatment, and three months after treatment. Psychiatric comorbidities were assessed using the Mini-International Neuropsychiatric Interview [29].

The primary clinical outcome was continuous treatment response, defined as percentage reduction in Y-BOCS total score from baseline to three-month follow-up. Participants with missing follow-up Y-BOCS scores were excluded (n = 19). For visualization and comparisons with controls, categorical response was defined as a reduction of at least 35% in Y-BOCS score, consistent with international consensus criteria. [30].

### Sample Collection and DNA Methylation Profiling

Saliva samples were collected using Oragene-DNA (OG500) kits (DNA Genotek Inc., Ottawa, Canada). Baseline and post-treatment samples were collected under supervision; follow-up samples were self-collected and mailed in. Controls provided saliva at a single time point.

DNA was extracted at *HUNT* (Norwegian University of Science and Technology, Trondheim, Norway) and *Life & Brain GmbH* (Bonn, Germany). DNA methylation was profiled in two phases (2023 and 2024) at *Life & Brain GmbH* using the Infinium MethylationEPIC v2.0 BeadChip array (*Illumina*, San Diego, USA). Sample placement on slides was randomized and balanced for sex, age, response status, and clinic site. Further details are described in the **Supplementary Materials**.

### Computational Framework

All preprocessing, quality control, statistical analyses and data visualization were conducted in R version 4.4.0.

### DNA Methylation Processing and Quality Control

DNA methylation data were preprocessed and quality-controlled using an adapted *CPACOR* [31] pipeline and annotated using hg38, as previously described [32]. Probes were excluded for: poor performance, cross-reactivity, common polymorphisms, mapping ambiguity, and sex-chromosome location. Samples were excluded for: sex mismatch, genotype discordance across repeated samples, poor bisulfite conversion, excess missingness, and technical outlier status.

### Statistical Analyses

Treatment response associations with demographic and clinical characteristics were examined using nonparametric or categorical tests, as appropriate.

Three epigenome-wide models were used:

1. **Baseline model:** association of pretreatment methylation with subsequent treatment response, tested using linear models with empirical Bayes moderation implemented in *limma* [33] (Figure 1A).
2. **Time-stable model:** association of repeated methylation measures with treatment response across time points, tested using linear mixed-effects models [34] (Figure 1B).
3. **Longitudinal change model:** interaction between treatment response and time point, testing differential within-person methylation change using *lmrse* [35] (Figure 1C).

Longitudinal analyses included participants with data from at least two time points. All models adjusted for age, sex, collection year, smoking exposure, technical covariates, ancestry, and cell-composition.

False discovery rate correction was applied. Quantile-quantile plots [36] and genomic inflation factors were examined (**Figure S1**).

Additional details are provided in the **Supplementary Materials**.

### Differentially Methylated Regions

Differentially methylated regions (DMRs) were identified using *ENmix-combp* [37,38] with a maximum probe distance of 750 bp and a seed p-value threshold of 0.05 [39]. Regions required ≥3 nominally significant probes, with >50% meeting nominal significance. Sidak correction was applied to account for multiple testing.

### Estimated Cell-Type Composition

Reference-based deconvolution using *HEpiDISH* [40] was used to estimate saliva cell-type proportions as previously described [32]. Associations between estimated cell-type proportions and treatment response were tested using baseline, time-stable, and longitudinal models analogous to the primary methylation analyses. Additional methodological details are provided in the **Supplementary Materials**.

### Secondary Analyses

Gene ontology enrichment analyses were performed using *missMethyl* [41]. Significant DMRs were examined for local methylation quantitative trait loci (mQTLs) in participants with genotype data (baseline: n = 371; stable: n = 353; longitudinal change: n = 352), and significant mQTL variants were subsequently tested for association with treatment response. Significant regions were also compared between responders, nonresponders, and healthy controls and evaluated in sensitivity analyses for medication use and psychiatric comorbidity. Cross-tissue relevance was assessed using an external saliva-brain methylation correlation dataset [42], with enrichment evaluated by permutation testing, as previously described [32]. Detailed procedures are reported in the **Supplementary Materials**.

## Results

### Sample Characteristics

After quality control, our sample comprised 889 individuals with OCD and 384 healthy controls (**Table S2**). Mean Y-BOCS scores decreased from 25.8 at baseline to 11.1 post-treatment and 10.1 at three-month follow-up (**Figure S2**). Lower treatment response was associated with older age, male sex, and mood stabilizer use. Comorbid depression, bulimia nervosa, and generalized anxiety disorder were associated with poorer response, whereas panic disorder was associated with greater improvement (**Table S3**).

### Baseline DNA Methylation Associations With Subsequent Treatment Response

Baseline analyses identified ten differentially methylated regions (DMRs) associated with subsequent treatment response after multiple-testing correction (Sidak-adjusted p < 0.05; **Table 1**, Figure 1A; Figure 2A-F, **Figure S3**). No individual methylation site reached epigenome-wide significance (**Table S4A**), and gene set enrichment analysis did not identify any significantly enriched biological processes (**Table S4B**).

**Figure 2:**
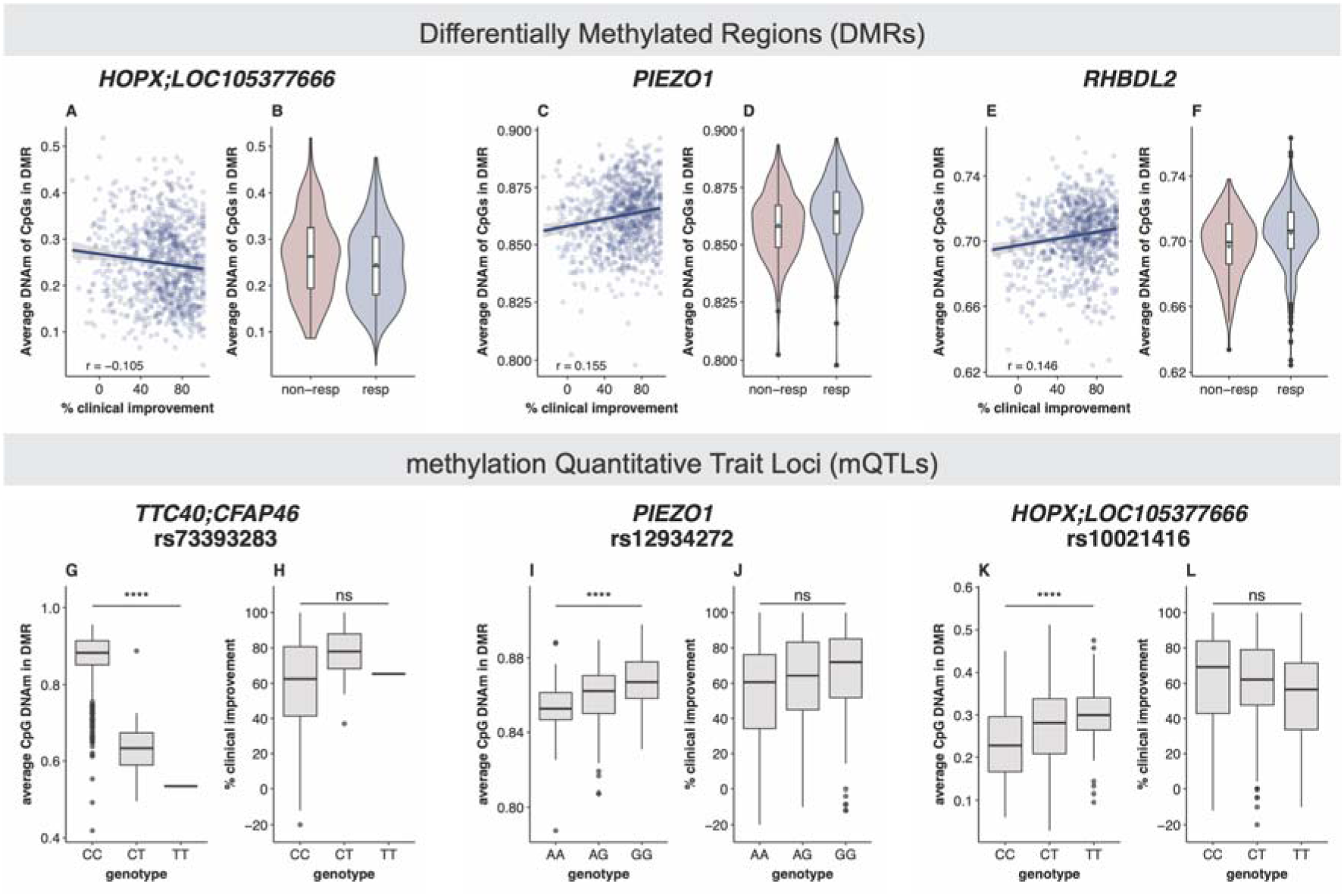
Examples of Baseline Differentially Methylated Regions (DMRs). A-F) DMR DNA Methylation associated with treatment response. G-L) Association between methylation Quantitative Trait Locus (mQTL) SNPs with DNA methylation in the respective DMR and treatment response. DNAm: DNA methylation (beta values), non-resp: non-responder, Pearson’s correlation coefficient, resp: responder, **** p < 0.0001.

**Table 1:**
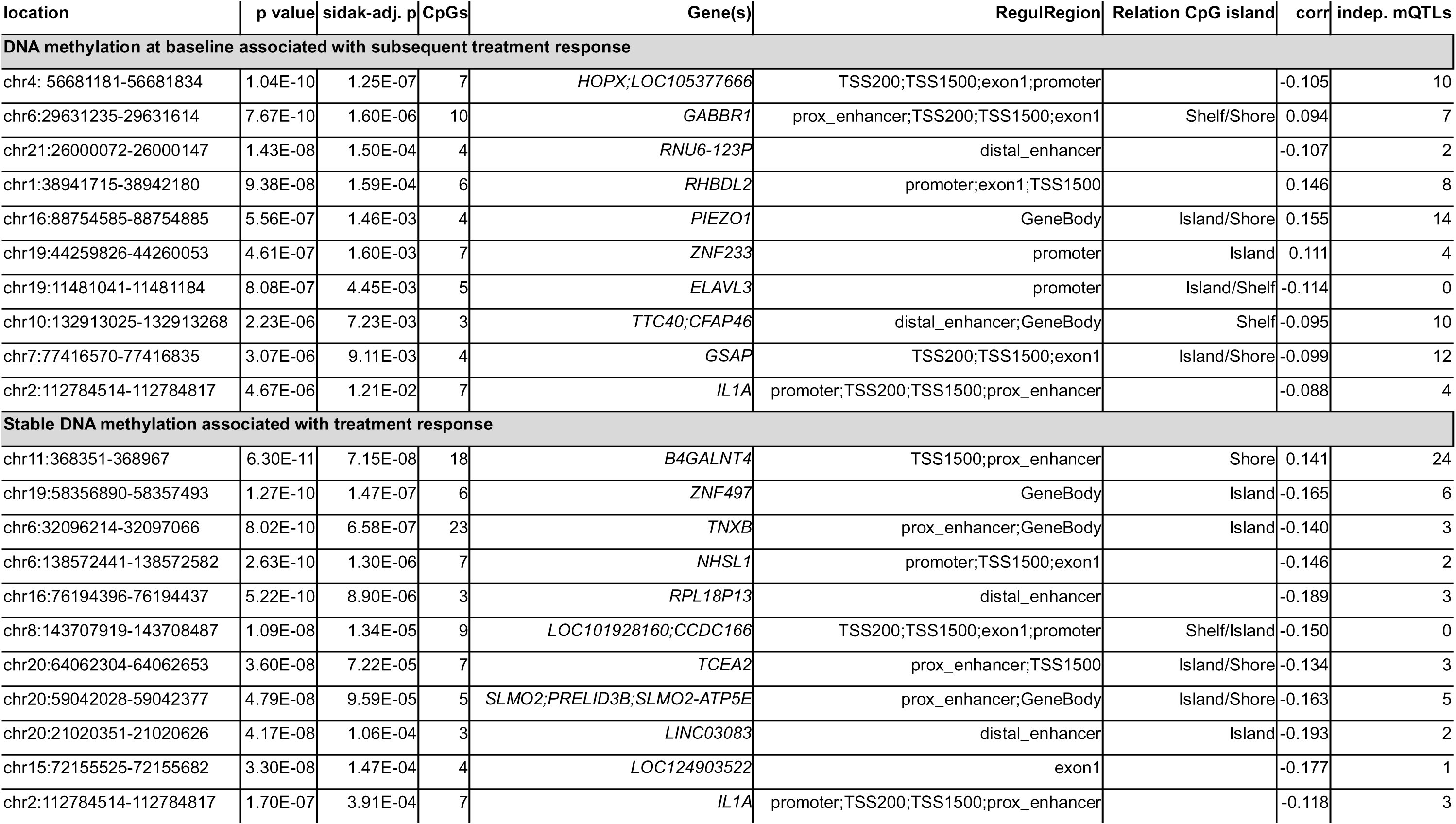
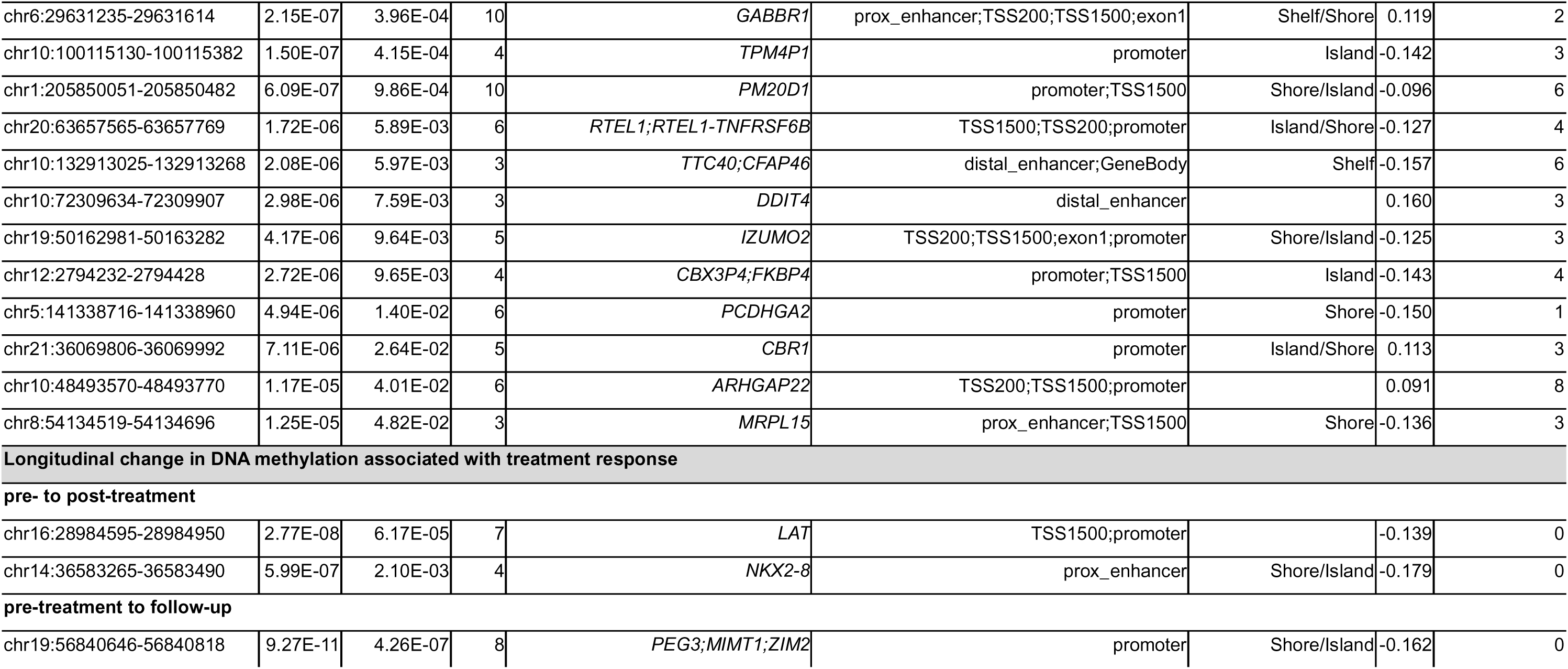
Differentially methylated regions (DMRs) associated with treatment response. Genomic location annotated using hg38. adj: adjusted, avg: average (here: across all time points and CpGs within the DMRs), chr: chromosome, corr: Pearson’s correlation coefficient (for the correlation between treatment response and average DNA methylation across CpGs for the respective DMR; for the stable over time analyses average DNA methylation across time time points; for the longitudinal change analyses change in DNA methylation), coefficient, indep.: independent after clumping, mQTL: methylation quantitative trait locus, RegulRegion: regulatory region, SE: standard error.

| location | p value | sidak-adj. p | CpGs | Gene(s) | RegulRegion | Relation CpG island | corr | indep. mQTLs |
| --- | --- | --- | --- | --- | --- | --- | --- | --- |
| **DNA methylation at baseline associated with subsequent treatment response** |  |  |  |  |  |  |  |  |
| chr4:56681181-56681834 | 1.04E-10 | 1.25E-07 | 7 | *HOPX;LOC105377666* | TSS200;TSS1500;exon1;promoter |  | -0.105 | 10 |
| chr6:29631235-29631614 | 7.67E-10 | 1.60E-06 | 10 | *GABBR1* | prox_enhancer;TSS200;TSS1500;exon1 | Shelf/Shore | 0.094 | 7 |
| chr21:26000072-26000147 | 1.43E-08 | 1.50E-04 | 4 | *RNU6-123P* | distal_enhancer |  | -0.107 | 2 |
| chr1:38941715-38942180 | 9.38E-08 | 1.59E-04 | 6 | *RHBDL2* | promoter;exon1;TSS1500 |  | 0.146 | 8 |
| chr16:88754585-88754885 | 5.56E-07 | 1.46E-03 | 4 | *PIEZO1* | GeneBody | Island/Shore | 0.155 | 14 |
| chr19:44259826-44260053 | 4.61E-07 | 1.60E-03 | 7 | *ZNF233* | promoter | Island | 0.111 | 4 |
| chr19:11481041-11481184 | 8.08E-07 | 4.45E-03 | 5 | *ELAVL3* | promoter | Island/Shelf | -0.114 | 0 |
| chr10:132913025-132913268 | 2.23E-06 | 7.23E-03 | 3 | *TTC40;CFAP46* | distal_enhancer;GeneBody | Shelf | -0.095 | 10 |
| chr7:77416570-77416835 | 3.07E-06 | 9.11E-03 | 4 | *GSAP* | TSS200;TSS1500;exon1 | Island/Shore | -0.099 | 12 |
| chr2:112784514-112784817 | 4.67E-06 | 1.21E-02 | 7 | *IL1A* | promoter;TSS200;TSS1500;prox_enhancer |  | -0.088 | 4 |
| **Stable DNA methylation associated with treatment response** |  |  |  |  |  |  |  |  |
| chr11:368351-368967 | 6.30E-11 | 7.15E-08 | 18 | *B4GALNT4* | TSS1500;prox_enhancer | Shore | 0.141 | 24 |
| chr19:58356890-58357493 | 1.27E-10 | 1.47E-07 | 6 | *ZNF497* | GeneBody | Island | -0.165 | 6 |
| chr6:32096214-32097066 | 8.02E-10 | 6.58E-07 | 23 | *TNXB* | prox_enhancer;GeneBody | Island | -0.140 | 3 |
| chr6:138572441-138572582 | 2.63E-10 | 1.30E-06 | 7 | *NHSL1* | promoter;TSS1500;exon1 |  | -0.146 | 2 |
| chr16:76194396-76194437 | 5.22E-10 | 8.90E-06 | 3 | *RPL18P13* | distal_enhancer |  | -0.189 | 3 |
| chr8:143707919-143708487 | 1.09E-08 | 1.34E-05 | 9 | *LOC101928160;CCDC166* | TSS200;TSS1500;exon1;promoter | Shelf/Island | -0.150 | 0 |
| chr20:64062304-64062653 | 3.60E-08 | 7.22E-05 | 7 | *TCEA2* | prox_enhancer;TSS1500 | Island/Shore | -0.134 | 3 |
| chr20:59042028-59042377 | 4.79E-08 | 9.59E-05 | 5 | *SLMO2;PRELID3B;SLMO2-ATP5E* | prox_enhancer;GeneBody | Island/Shore | -0.163 | 5 |
| chr20:21020351-21020626 | 4.17E-08 | 1.06E-04 | 3 | *LINC03083* | distal_enhancer | Island | -0.193 | 2 |
| chr15:72155525-72155682 | 3.30E-08 | 1.47E-04 | 4 | *LOC124903522* | exon1 |  | -0.177 | 1 |
| chr2:112784514-112784817 | 1.70E-07 | 3.91E-04 | 7 | *IL1A* | promoter;TSS200;TSS1500;prox_enhancer |  | -0.118 | 3 |
| chr6:29631235-29631614 | 2.15E-07 | 3.96E-04 | 10 | *GABBR1* | prox_enhancer;TSS200;TSS1500;exon1 | Shelf/Shore | 0.119 | 2 |
| chr10:100115130-100115382 | 1.50E-07 | 4.15E-04 | 4 | *TPM4P1* | promoter | Island | -0.142 | 3 |
| chr1:205850051-205850482 | 6.09E-07 | 9.86E-04 | 10 | *PM20D1* | promoter;TSS1500 | Shore/Island | -0.096 | 6 |
| chr20:63657565-63657769 | 1.72E-06 | 5.89E-03 | 6 | *RTEL1;RTEL1-TNFRSF6B* | TSS1500;TSS200;promoter | Island/Shore | -0.127 | 4 |
| chr10:132913025-132913268 | 2.08E-06 | 5.97E-03 | 3 | *TTC40;CFAP46* | distal_enhancer;GeneBody | Shelf | -0.157 | 6 |
| chr10:72309634-72309907 | 2.98E-06 | 7.59E-03 | 3 | *DDIT4* | distal_enhancer |  | 0.160 | 3 |
| chr19:50162981-50163282 | 4.17E-06 | 9.64E-03 | 5 | *IZUMO2* | TSS200;TSS1500;exon1;promoter | Shore/Island | -0.125 | 3 |
| chr12:2794232-2794428 | 2.72E-06 | 9.65E-03 | 4 | *CBX3P4;FKBP4* | promoter;TSS1500 | Island | -0.143 | 4 |
| chr5:141338716-141338960 | 4.94E-06 | 1.40E-02 | 6 | *PCDHGA2* | promoter | Shore | -0.150 | 1 |
| chr21:36069806-36069992 | 7.11E-06 | 2.64E-02 | 5 | *CBR1* | promoter | Island/Shore | 0.113 | 3 |
| chr10:48493570-48493770 | 1.17E-05 | 4.01E-02 | 6 | *ARHGAP22* | TSS200;TSS1500;promoter |  | 0.091 | 8 |
| chr8:54134519-54134696 | 1.25E-05 | 4.82E-02 | 3 | *MRPL15* | prox_enhancer;TSS1500 | Shore | -0.136 | 3 |
| pre- to post-treatment |  |  |  |  |  |  |  |  |
| chr16:28984595-28984950 | 2.77E-08 | 6.17E-05 | 7 | *LAT* | TSS1500;promoter |  | -0.139 | 0 |
| chr14:36583265-36583490 | 5.99E-07 | 2.10E-03 | 4 | *NKX2-8* | prox_enhancer | Shore/Island | -0.179 | 0 |
| pre-treatment to follow-up |  |  |  |  |  |  |  |  |
| chr19:56840646-56840818 | 9.27E-11 | 4.26E-07 | 8 | *PEG3;MIMT1;ZIM2* | promoter | Shore/Island | -0.162 | 0 |

Nine of the ten baseline DMRs showed evidence of local methylation quantitative trait loci (mQTLs), indicating substantial genetic regulation (**Table 2, Table S4C-D)**. However, associated variants were not significantly related to treatment response after correction for multiple testing (**Table S4D**, Figure 2G-L).

In case-control comparisons, DNA methylation at the *GABBR1* DMR differed between healthy controls and both responders (q = 0.0164) and nonresponders (q = 0.0009), whereas the *RHBDL2* DMR differed primarily in nonresponders (q = 0.0004; **Table S4E**).

Sensitivity analyses suggested limited effects of medication use or psychiatric comorbidity on baseline findings (**Table S4F-G**), with a significant effect only for psychoactive medication at the *RNU6-123P* DMR (q = 0.046).

### Time-Stable DNA Methylation Associations With Treatment Response

Across time points, 23 DMRs showed stable associations between DNA methylation and treatment response (Sidak-adjusted p < 0.05; **Table 1**, Figure 1B; Figure 3A-D**, Figure S4**). Three overlapped with baseline-associated DMR (*GABBR1, TTC40/CFAP46,* and *IL1A*). No single methylation site reached significance after multiple-testing correction (FDR-adjusted p < 0.05; **Table S5A**). Gene set enrichment analysis did not identify any significantly enriched biological processes; however, terms related to neurogenesis and neuronal function were among the top-ranked categories (**Table S5B**).

**Figure 3:**
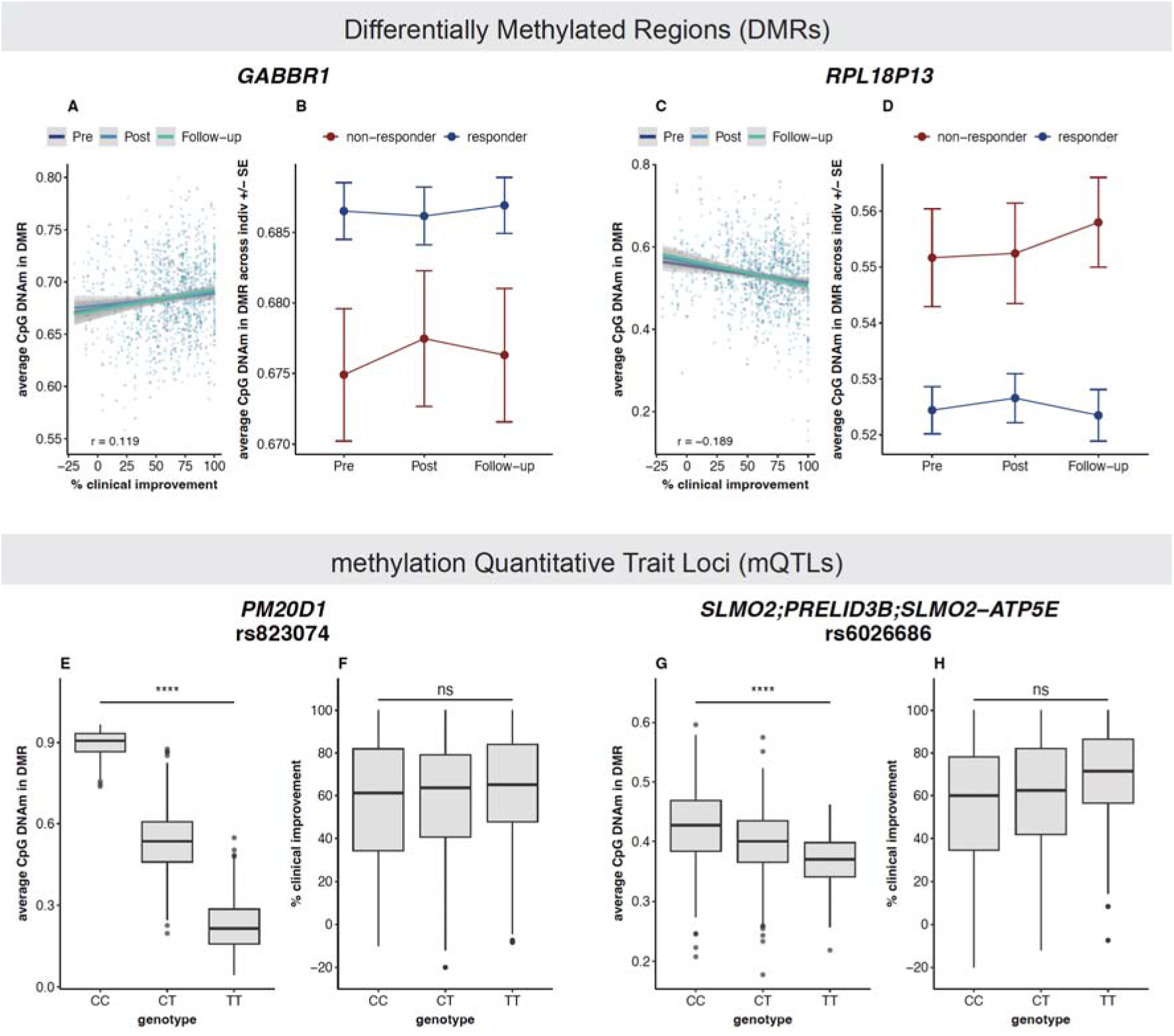
Examples of Stable Differentially Methylated Regions (DMRs). A-D) DMR DNA Methylation associated with treatment response across different time points. E-F) Association between methylation Quantitative Trait Locus (mQTL) SNPs with DNA methylation in the DMR and with treatment response. DNAm: DNA methylation (beta values), non-resp: non-responder, Pearson’s correlation coefficient, resp: responder, **** p < 0.0001.

Twenty-two of the 23 stable DMRs showed significant local genetic regulation (**Table S5C-D**). When testing the association between independent mQTLs and treatment response, thirteen showed nominal associations (Figure 3E-H**, Table S5D**); however, none remained significant after multiple-testing correction.

Compared with healthy controls, non-responders generally showed greater methylation differences than responders across several regions (**Table S5E**). The DMR annotated to *GABBR1* showed significant differences between controls and responders, as well as between controls and non-responders, at all time points.

Some time-stable associations were influenced by medication use or specific psychiatric comorbidities in sensitivity analyses, suggesting partial clinical confounding (**Table S5F-G**).

### Longitudinal DNA Methylation Change Association With Treatment Response

Longitudinal analyses identified three DMRs in which within-person methylation change was associated with treatment response: *LAT* and *NKX2-8* from pretreatment to posttreatment, and *PEG3/MIMT1/ZIM2* from pretreatment to follow-up (Sidak-adjusted p < 0.05; **Table 1**, Figure 1C**, Figures S5-S7**). No single methylation site reached epigenome-wide significance (**Table S6A**). No biological processes were significantly enriched after multiple-testing correction, although nervous system development ranked among the top categories in the pre-post analysis (**Table S6B**).

No significant mQTLs were detected for these longitudinal change signals. Associations were more sensitive to anxiolytic medication use and bulimia nervosa comorbidity, suggesting greater vulnerability to time-varying clinical factors (**Table S6C-D**).

### Cross-Tissue Correlation Analyses

Several CpG sites within significant DMRs showed nominal saliva-brain methylation correlations in an external reference dataset [42] (**Table S4H, Table S5H, Table S6E**). Enrichment of cross-tissue correlation was strongest for stable regions: four DMRs, annotated to *ZNF497*, *LOC101928160/CCDC166*, *PM20D1*, and *IZUMO2*, showed significant enrichment for saliva-brain correlation in permutation tests, although most regions did not show robust enrichment (**Table S5H**).

### Estimated Cell-Type Composition

Estimated saliva cell-type proportions varied substantially across samples (**Figure S8**) and changed over time for several cell types (**Figure S9**, **Table S7A**). Greater treatment response was weakly associated with changes in monocyte proportions from pretreatment to posttreatment (p = 0.0276) and with CD4+ T-cell changes from pretreatment to follow-up (p = 0.0471; **Table S7B**). No significant associations were observed for baseline cell-type proportions or time-stable models (**Table S7A**,**C**).

Sensitivity analyses indicated that these associations were influenced by medication and comorbidities (**Table S7D-E**). Overall, smoking exposure and age explained substantially more variance in estimated cell type proportions than treatment response (**Table S7A,C**).

## Discussion

In this longitudinal EWAS of 889 adults with OCD receiving intensive exposure and response prevention psychotherapy, we identified three classes of DNA methylation signals associated with treatment response: ten pretreatment differentially methylated regions (DMRs) associated with later response, 23 time-stable DMRs showing consistent associations across repeated assessments, and three dynamic DMRs in which methylation change correlated with clinical improvement. Baseline and time-stable findings were frequently influenced by nearby genetic variation, whereas longitudinal signals appeared less genetically influenced and more sensitive to clinical factors such as medication use and psychiatric comorbidity. Together, these results suggest that psychotherapy response may reflect both trait-like and state-dependent biological processes.

Several identified DMRs were annotated to genes involved in neurotransmission, neuroplasticity, and stress-related signaling, processes relevant to extinction learning [43], which is central to successful exposure-based therapy [44]. Of particular interest, *GABBR1* was associated with treatment response in both baseline and time-stable analyses and also differed from healthy controls. *GABBR1* encodes a subunit of the GABA_B_ receptor and has shown suggestive associations with treatment response in an epigenetic study of children and adolescents with OCD (54), and has also been implicated in OCD in prior genetic and epigenetic studies, including a case-control EWAS in a subset of this cohort [32,46,47]. Together, these findings support a potential role for inhibitory signaling in treatment responsiveness [48].

Additional DMRs were annotated to genes involved in neurodevelopment, synaptic organization, neuronal connectivity, and extracellular matrix biology, including *HOPX, PIEZO1, ELAVL3, PCDHGA2, NHSL1, ARHGAP22,* and *TNXB* [49–60]. *TNXB* has also been identified in OCD and other neuropsychiatric disorders in prior epigenetic studies [47,61–64]. Although pathway analyses did not survive multiple testing correction, they were broadly consistent with enrichment of neurodevelopmental and neuronal regulatory processes.

We also observed associations in genes related to immune regulation and cellular metabolism, including *IL1A, LAT, MRPL15, SLMO2/PRELID3B, DDIT4, and PEG3*. These findings align with growing evidence implicating inflammatory and metabolic pathways in OCD [65,66]. Modest associations between treatment response and changes in estimated monocyte and CD4+ T-cell proportions further support the possibility that peripheral immune processes may accompany clinical improvement in a subset of patients, although these effects were small and influenced by medication use and comorbidity.

A substantial proportion of identified loci mapped to genes involved in transcriptional and post-transcriptional regulation, including zinc-finger transcription factors, non-coding RNAs, and other regulatory elements. This pattern suggests that treatment response-associated methylation signals may partly influence downstream molecular pathways through altered gene regulation.

Apart from *GABBR1*, we did not detect differential methylation in candidate genes previously implicated in psychotherapy response. This discrepancy may reflect differences in tissue type, sample size, intervention specificity, and the limited reproducibility of earlier candidate-gene results in epigenome-wide studies.

Gene-level findings should be interpreted cautiously. The present results are based on peripheral saliva DNA methylation and reflect associations rather than direct evidence of causal brain mechanisms. Several DMRs, including regions annotated to *ZNF497, LOC101928160/CCDC166, PM20D1,* and *IZUMO2*, showed enrichment of saliva-brain methylation correlations, suggesting that some peripheral signals may reflect biologically relevant central processes. However, brain tissue was not assessed directly, and the reference dataset was limited in size and heterogenous in brain-region coverage [42]. Even when saliva does not closely mirror brain methylation, it may still reflect systemic processes such as inflammation or stress regulation and serve as a clinically informative biomarker of treatment response.

An important finding was the extensive local genetic regulation of baseline and time-stable methylation signals. Most significant regions in these analyses showed nearby methylation quantitative trait loci (mQTLs), suggesting that stable methylation differences associated with psychotherapy outcome may partly reflect inherited biological liability. However, no associated variants were directly related to treatment response after multiple-testing correction, consistent with the small effect sizes expected for complex clinical phenotypes [67]. By contrast, none of the longitudinal change signals showed significant mQTL effects. This pattern is consistent with the interpretation that dynamic methylation changes may be more responsive to environmental or treatment-related influences, whereas stable signatures may reflect pre-existing susceptibility or treatment capacity.

The distinction between baseline, time-stable, and dynamic associations may be clinically relevant. Pretreatment methylation signals could eventually contribute to stratification approaches that identify individuals less likely to respond to standard exposure-based therapy. Time-stable signals may reflect trait-like biological characteristics associated with treatment capacity, whereas dynamic signals may capture state-related processes accompanying symptom change, stress reactivity, epigenetic instability, or heterogeneity in environmental exposures.

### Strengths and Limitations

This study has several strengths. To our knowledge, it is the largest epigenomic investigation of response to psychological treatment and the first to integrate mQTL analyses in this context. The longitudinal design enabled evaluation of both stable and dynamic methylation patterns, while the standardized treatment delivered in routine clinical settings enhances real-world relevance. In addition, the short treatment duration (four days) reduces the likelihood that observed methylation changes reflect lifestyle-related influences or natural temporal variation.

Several limitations should also be considered. First, despite being the largest study of its kind to date, power remained limited for single-site epigenome-wide discovery, and significant findings were stronger at the regional than individual CpG level. Longitudinal findings were fewer than anticipated, likely because interaction models generally require considerably larger sample sizes[68]. Second, the cohort showed high overall response rates, reducing outcome variability and potentially limiting statistical power to detect associations. Third, residual confounding by medication use, psychiatric comorbidity, smoking, or unmeasured environmental exposures cannot be excluded, particularly for longitudinal analyses. Fourth, the sample was recruited in Norway and consisted predominantly of individuals of European ancestry, which may limit generalizability.

### Outlook

Future research should replicate these findings in independent and more diverse cohorts and examine whether the observed methylation signatures generalize across different psychotherapeutic and pharmacological treatments. Integrating DNA methylation with clinical and genetic data may ultimately advance personalized treatment approaches in OCD.

## Conclusion

In conclusion, response to exposure-based psychotherapy in OCD was associated with both stable and dynamic peripheral DNA methylation patterns annotated to genes involved in neuroplasticity, stress response, immune function, mitochondrial processes, and gene regulation. Baseline and time-stable signals were frequently genetically regulated, whereas longitudinal changes appeared more influenced by clinical factors. These findings support DNA methylation as a promising biomarker of treatment response and may help clarify the biological mechanisms underlying therapeutic change.

## Data Availability

Due to Norwegian regulations, genetic and epigenetic data cannot be made publicly available. Summary statistics are available from the corresponding author upon request.

## Acknowledgements

We thank all study participants for their contribution. We are especially grateful to Helene Nilsen, Marie Bjorøy, Lisa Vårdal, Nadine Fricker at Life & Brain, and the participating psychotherapists for their roles in data collection and sample processing. We also thank Darina Czamara, Vera Karlbauer, Seyma Katrinli Wise, Alicia Schowe, Alicia Smith, Natan Yusupov, Torsten Klengel, and Lea Zillich for their helpful discussions on the use of the EPIC v2 array and longitudinal DNA methylation analyses. We used ChatGPT-5.3 (OpenAI, San Francisco, United States) to improve the clarity and style of the text; the scientific content was not altered.

## Author Contributions

SLH, KDH, ALT, OTO, GK, JJC, JH, KJR, and BH contributed to conceptualization of the study. TOE and KH were responsible for data collection. KDH conducted the epigenetic analyses and wrote the original draft of the manuscript. MWH conducted the genetic analyses. Supervision was provided by SLH, AKS, and KJR. Funding was acquired by ALT, OTO, GK, JJC, JH, KJR, BH, and SLH. All authors reviewed, revised, and approved the final manuscript.

