## Supplementary Materials for "Epigenetic Markers of Response to Psychotherapy in Obsessive-Compulsive Disorder"

#### Clinical Analyses

Case-control group differences in age and sex were assessed using Wilcoxon rank-sum and chi-square tests, respectively. Associations between treatment response and demographic or clinical variables were tested using Kendall’s tau for continuous variables and Wilcoxon rank-sum tests for categorical variables.

#### Profiling Phases

In phase I, individuals with OCD who provided all three samples were included. To complete array slides, matched control samples were placed on the same slides as their corresponding OCD cases; when a matched control was unavailable, samples from OCD participants with only two time points were used instead. In phase II, only pre-treatment samples from individuals with OCD were analyzed.

For the baseline analysis, Phase I and Phase II samples were combined because (1) they represent the same population without clinical differences (with balanced responder/non-responder distribution across phases); (2) samples were processed by the same laboratory; (3) control probe PCs together with collection year almost fully accounted for variation in phase; and (4) no inflation of p-values was observed when combining, indicating an absence of major batch effects.

#### Epigenome-Wide Association Analyses

DNA methylation associated with treatment response was identified using the following models:

1) Association between baseline DNA methylation and subsequent treatment response (**Figure 1A**):

M values at baseline ~ response + covariates,

2) Association between stable DNA methylation over time and treatment response, with response as the main effect and time point as a covariate (using *lmerTest* (1)):

M values ~ response + time point + covariates + (1 | individual ID),

3) Longitudinal DNA methylation change associated with treatment response (using *lmrse* (2)):

M values ~ %improvement × time point + covariates, cluster = individual ID

Probes showing singularity in the time-stable linear mixed effects model, due to limited variability or unstable estimates, were excluded. A more complex random intercept and slope model for longitudinal change was attempted but was not feasible given the high dimensionality of the data.

Covariates included sex, age, collection year, smoking exposure quantified as the M value of cg05575921 in *AHRR* (3,4), two principal components (PCs) of estimated cell-type proportions, control probe PCs to account for technical variation (5) (5 PCs in the baseline and longitudinal change analyses; 15 PCs in the time stable analysis, excluding control probe PC4 because of multicollinearity with sex), and one ancestry PC (6).

#### Functional Enrichment Analyses

Gene ontology analyses of biological processes were performed using goMeth() from the *missMethyl* package (7) on the top 0.5% of CpGs. P values were FDR-adjusted.

#### Genetic Analyses

##### Methylation Quantitative Trait Locus (mQTL) Analyses

In OCD participants with available genotyping data (baseline: n = 371; stable: n = 353; longitudinal change: n = 352), mQTL analyses were performed for each DMR by testing SNPs within ±50 kb for association with mean DMR methylation (see (8) for methodological details). Models matched those used in the main analyses: linear regression for baseline, linear mixed effects models for stable analyses, and linear regression with clustered robust standard errors for longitudinal change. All models were adjusted for the same covariates, except that 10 genotyping principal components were included instead of the ancestry PC. For the longitudinal change analysis, clustered standard errors were implemented using vcovCL() from the *sandwich* package (9) and coeftest() from the *lmtest* package (10). FDR correction was applied across all SNPs within each analysis.

##### Response Association of Significant mQTLs

For significant mQTLs, associations between the SNP (predictor) and treatment response (outcome) were tested using linear regression adjusted for age and ten genotyping PCs. Linkage disequilibrium clumping was performed in PLINK (11), retaining the top response-associated SNP per LD block. FDR correction was applied to independent mQTLs.

#### Comparison With Healthy Controls

For significant DMRs identified in the baseline and time-stable analyses, mean methylation levels were compared between responders, non-responders, and healthy controls using linear models adjusted for the same covariates as the main analyses, except collection year. Only samples from slides on which both cases and controls were typed together were included. For time-stable analyses, comparisons were performed separately for each time point. P values were FDR-adjusted.

#### Cell Type Proportion Analyses

Associations between estimated saliva cell-type proportions and treatment response were analyzed using baseline, time-stable, and longitudinal models analogous to the DNA methylation analyses. Models were adjusted for sex, age, smoking exposure, collection year, five control probe PCs (5), and one ancestry PC (6). To account for multiple testing, the Li and Ji method (12) was applied by multiplying p values by the effective number of independent tests (n = 1.85).

#### Sensitivity Analyses

The influence of psychiatric comorbidities and psychoactive medication classes on DMR results (average beta values across CpGs, transformed to M values) was evaluated by adding them as interaction terms in the main models. P-values within each comorbidity and medication class were FDR-adjusted.

Sensitivity analyses for the cell type proportion analyses mirrored those for DNA methylation, incorporating interactions with comorbidities and medication classes, and were applied to cell type proportions previously found to be significantly associated with treatment response.

#### Visualization of DNA Methylation Results

For visualization, M values were adjusted for the covariables included in the main models using *limma*’s removeBatchEffect function (13), then converted to beta values for plotting.

### Supplementary Figures

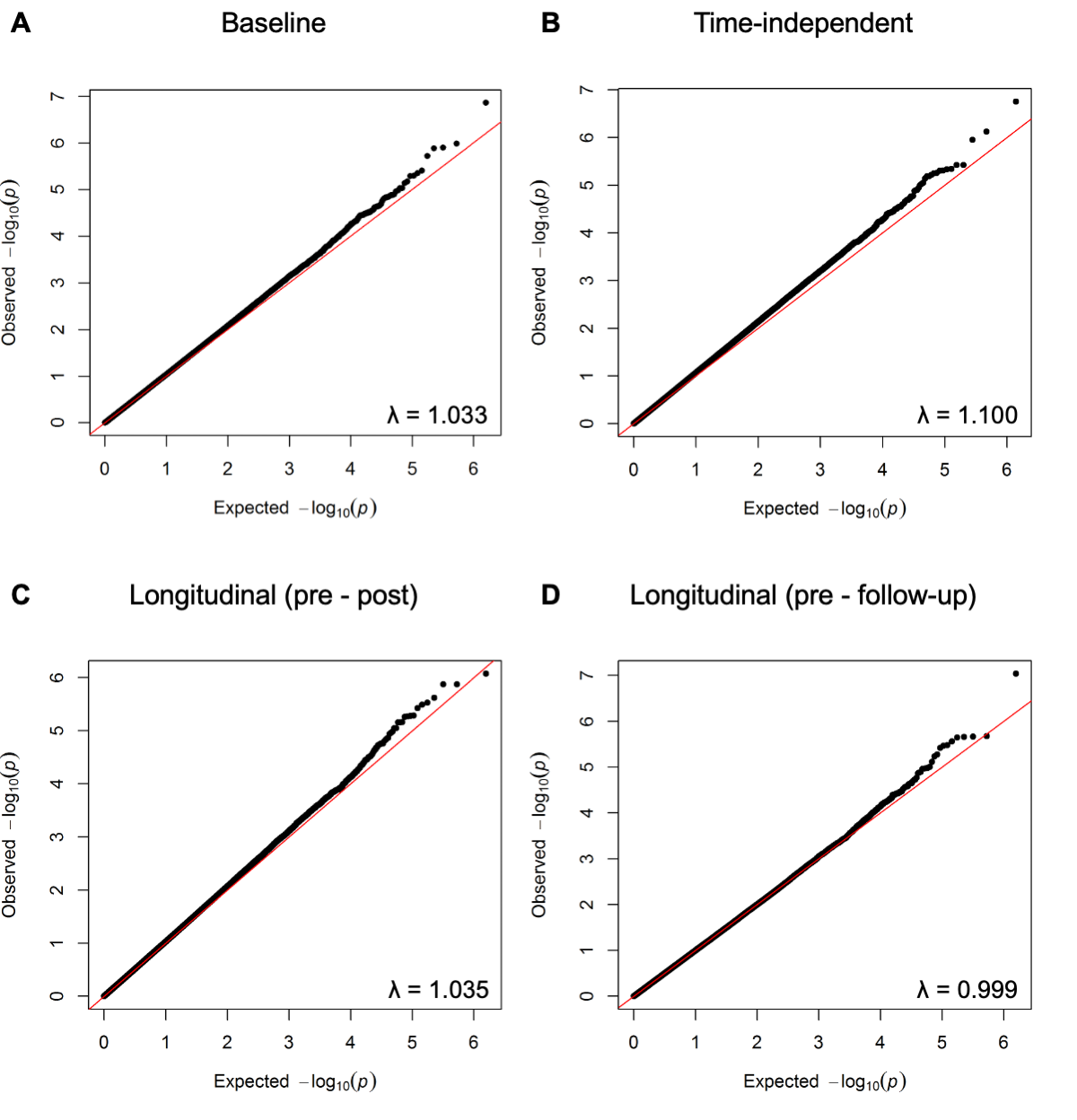

**Figure S1: Quantile-Quantile (QQ)-plots with genomic inflation factor (λ).**

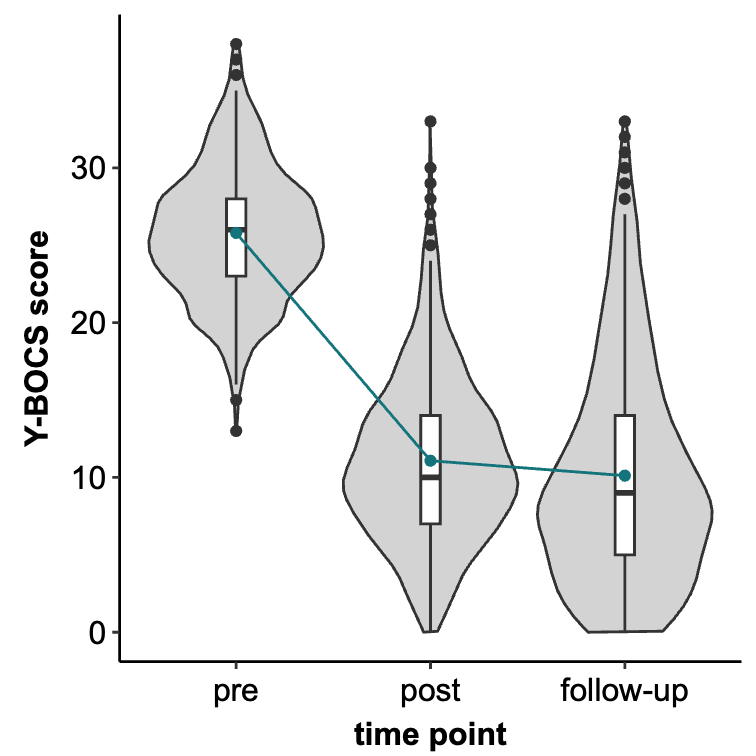

**Figure S2: Clinical improvement over the treatment and follow-up period.** Data were collected at three time points: pre-treatment, post-treatment (day 10), and follow-up (three months post-treatment). Y-BOCS: Yale–Brown Obsessive–Compulsive Scale.

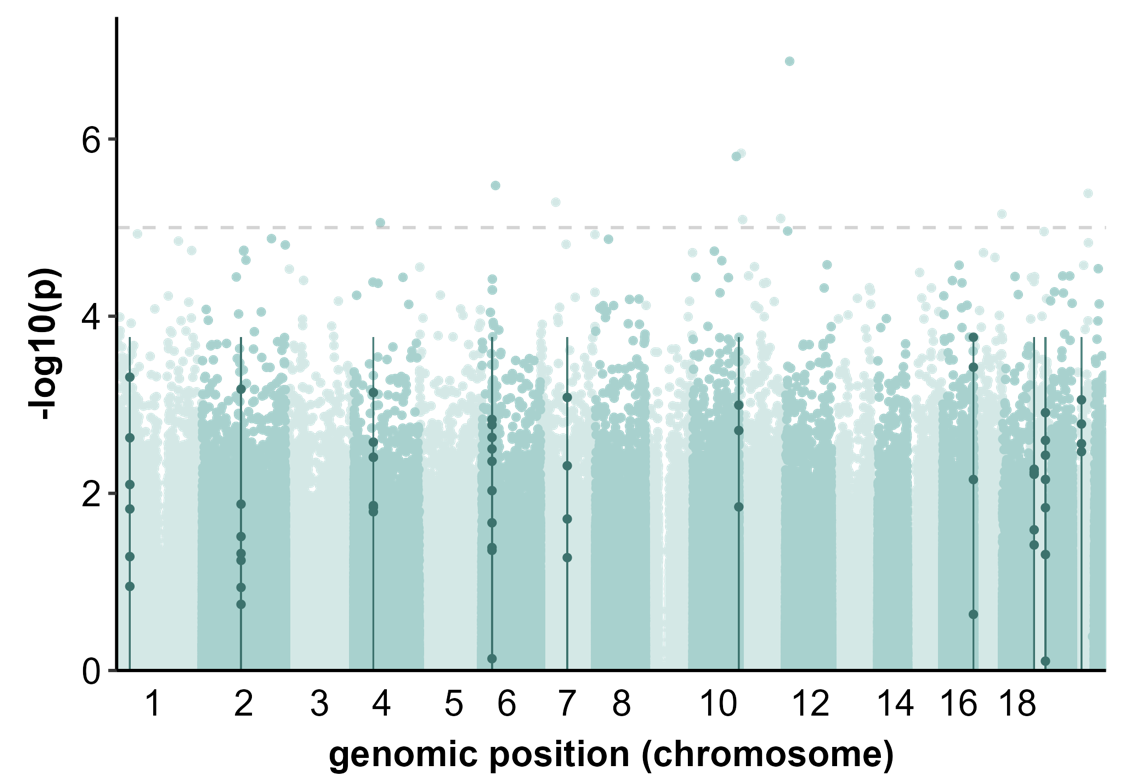

**Figure S3: Manhattan plot for the association between baseline DNA methylation and treatment response.**

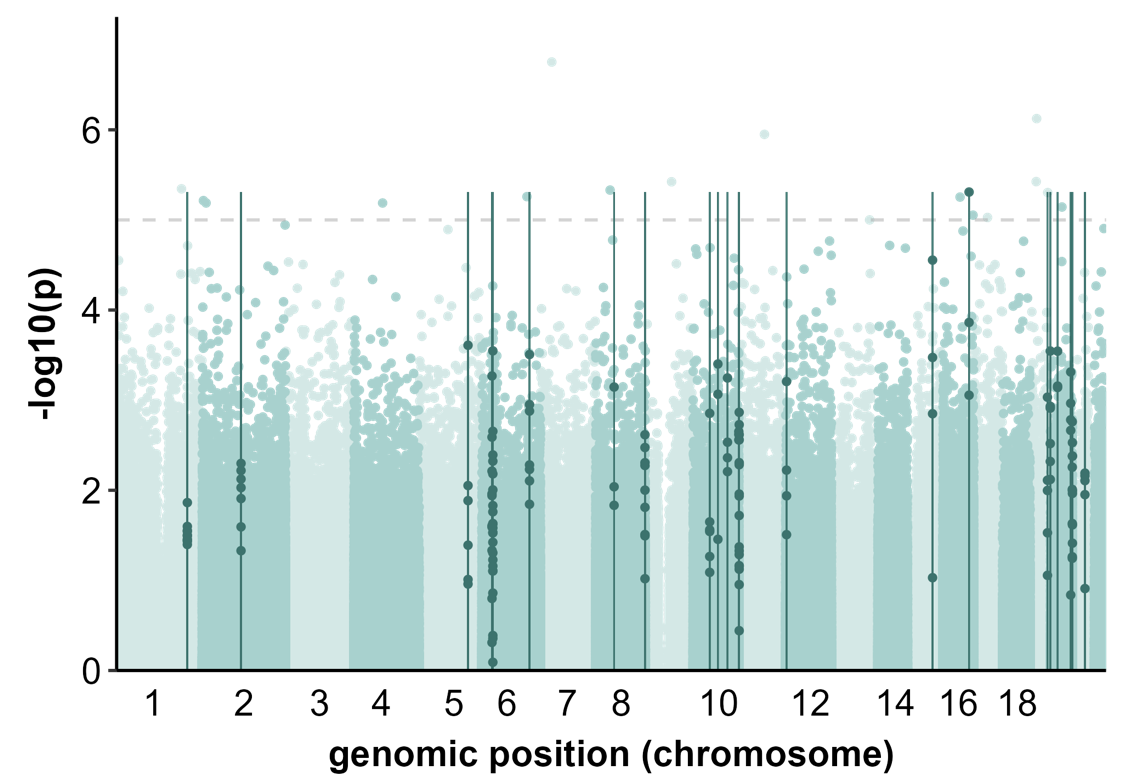

**Figure S4: Manhattan plot for the association between stable DNA methylation and treatment response.**

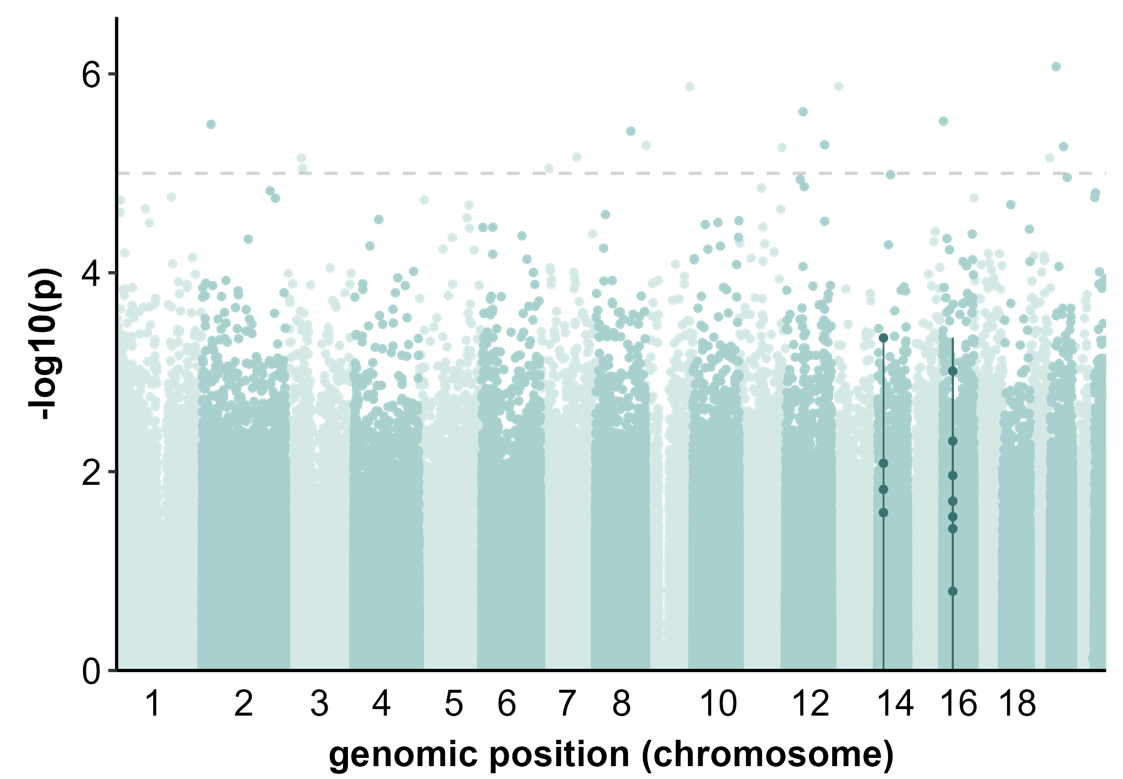

**Figure S5: Manhattan plot for the association between DNA methylation change from pre- to post-treatment and treatment response.**

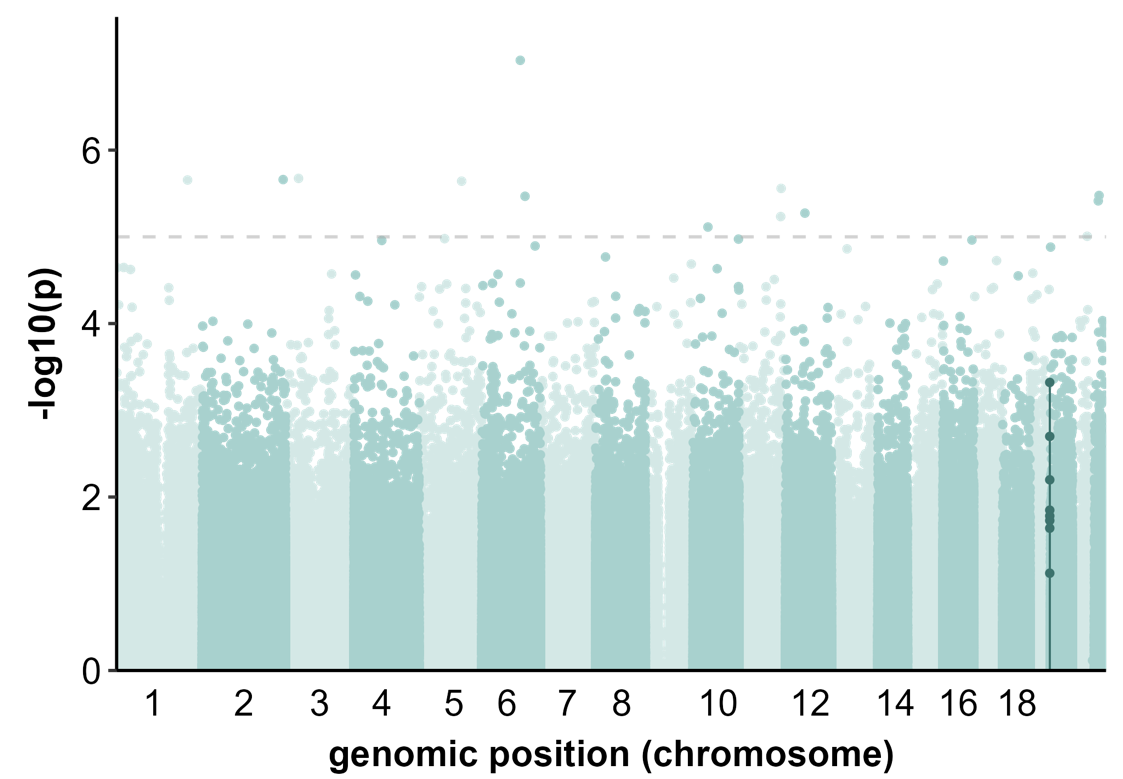

**Figure S6: Manhattan plot for the association between DNA methylation change from pre-treatment to the three-month follow-up and treatment response.**

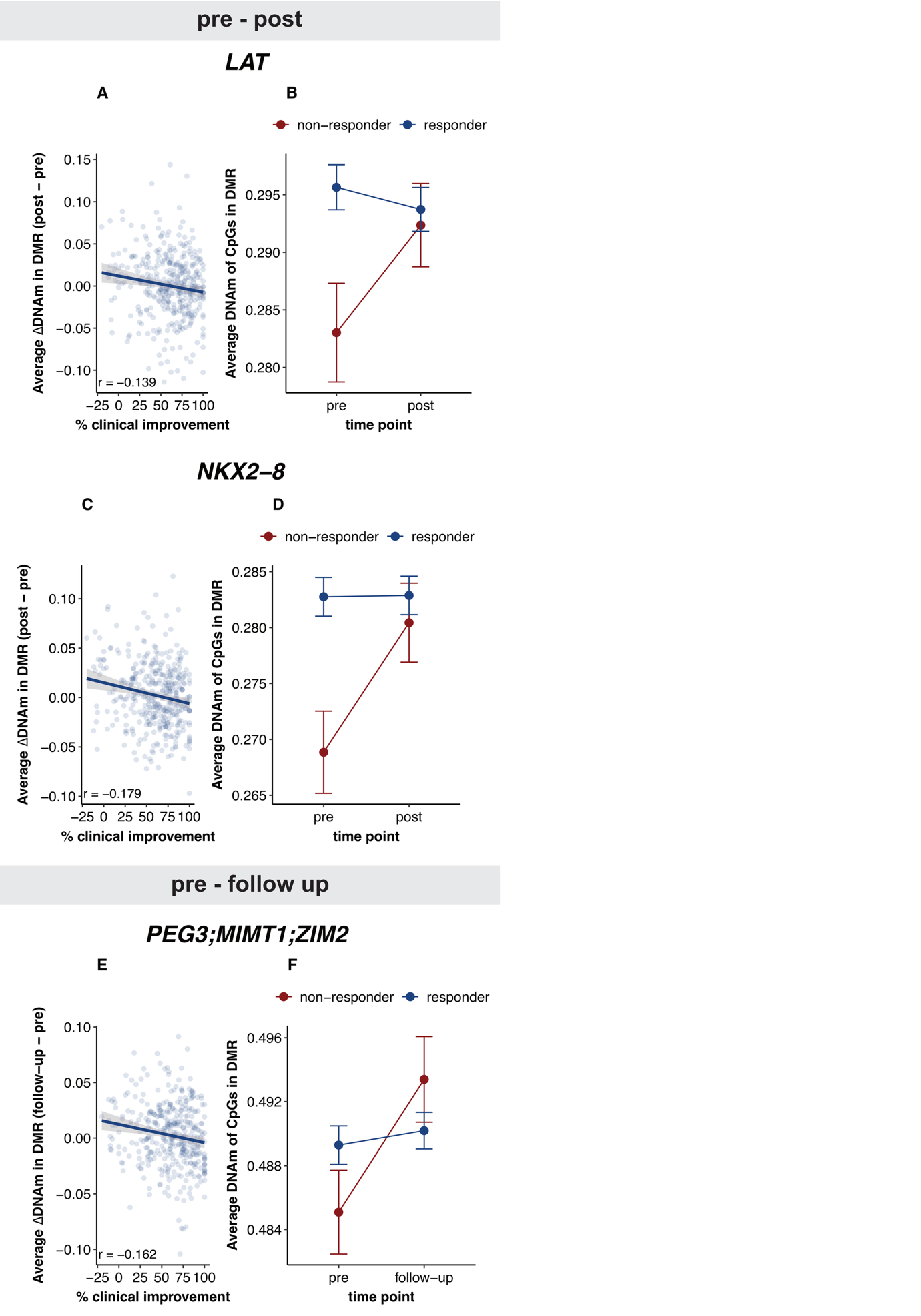

**Figure S7: Longitudinal DMRs associated with treatment response.** chr: chromosome, DMR: differentially methylated region, DNAm: DNA methylation, r: Pearson correlation coefficient.

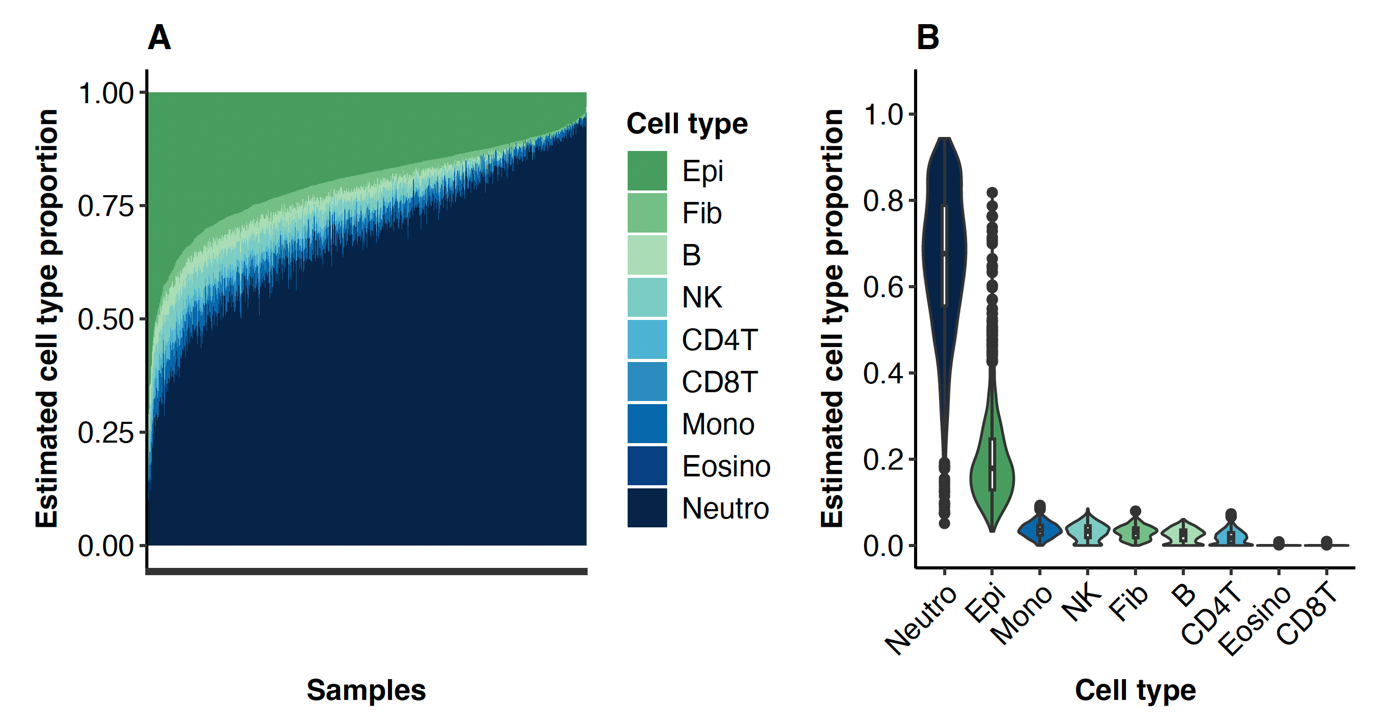

**Figure S8: Variation of estimated cell type proportions across all samples. B**: B cells, CD4T: CD4+ T cells, CD8T: CD8+ T cells, Epi: Epithelial cells, Eosino: Eosinophils, Fib: Fibroblasts, Mono: Monocytes, Neutro: Neutrophils, NK: Natural Killer cells.

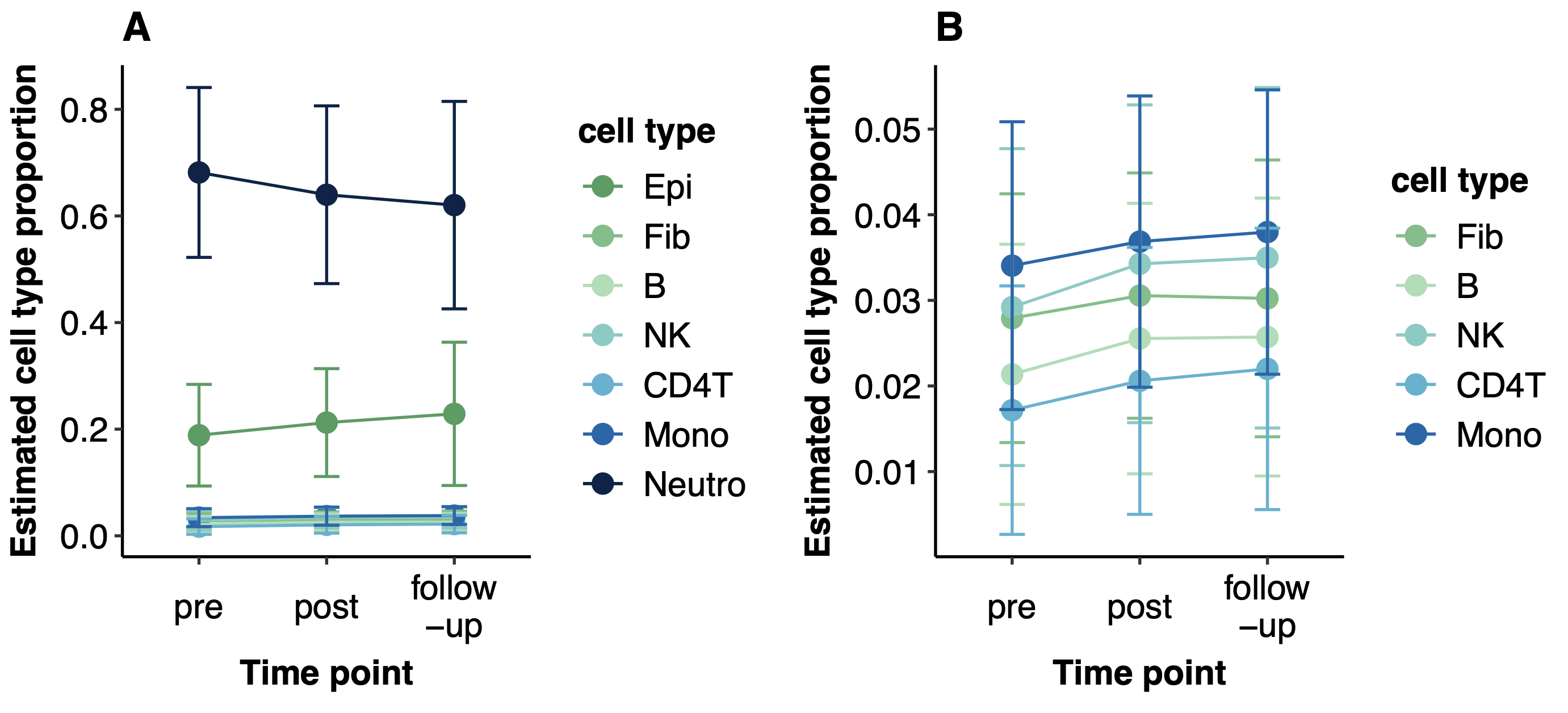

**Figure S9: Longitudinal change in estimated cell type proportions over time. A) All detectable cell type proportions, B) Zoomed in on estimated cell type proportions <0.1.** Plots show mean and standard deviation. B: B cells, CD4T: CD4+ T cells, Epi: Epithelial cells, Fib: Fibroblasts, Mono: Monocytes, Neutro: Neutrophils, NK: Natural Killer cells.

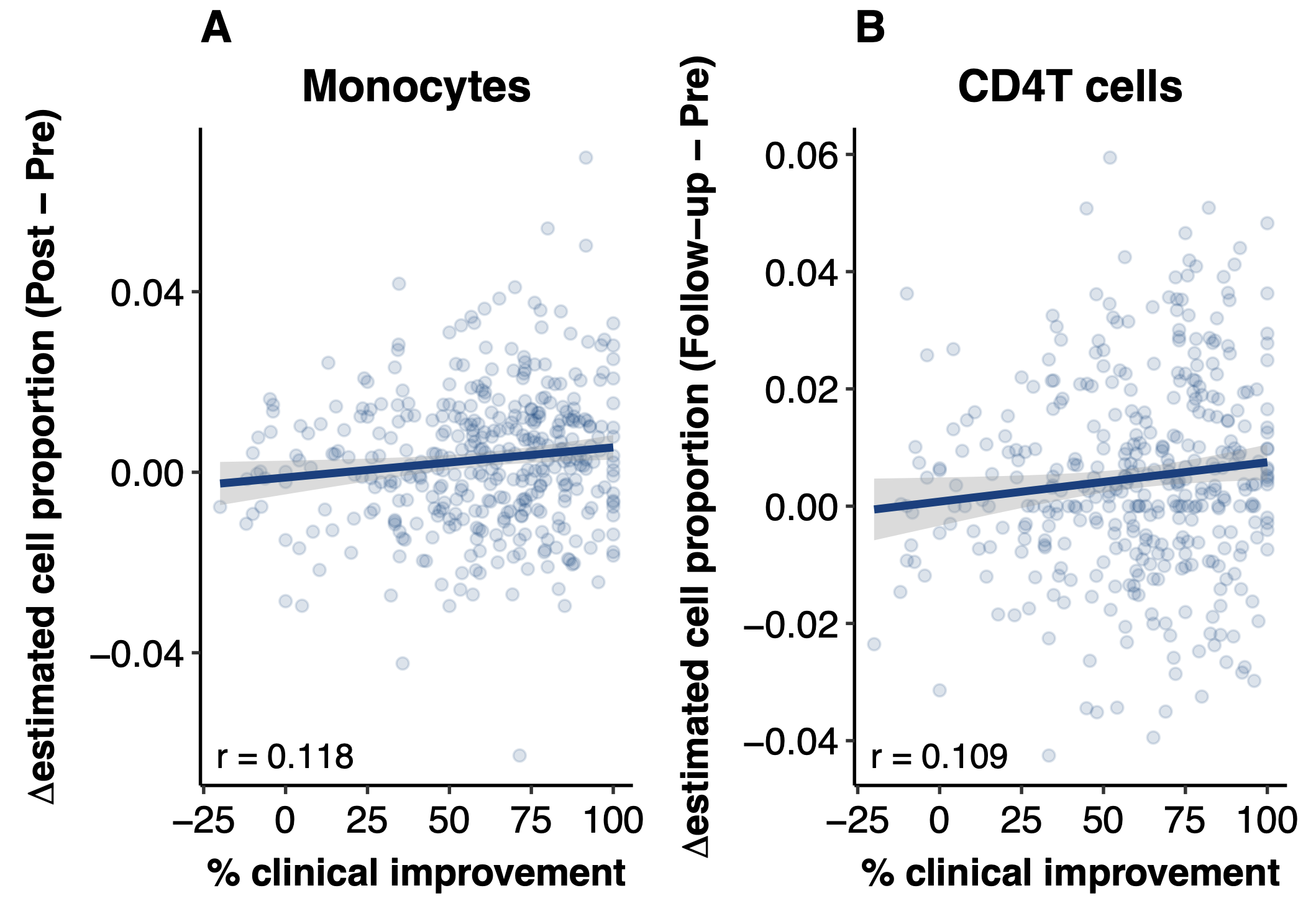

**Figure S10: Change in estimated cell type proportions significantly associated with response.** r: Pearson's correlation coefficient.

### Supplementary Tables

**Table S1: Psychoactive medication use among participants.** NaSSA: Noradrenergic and Specific Serotonergic Antidepressant, NDRI: Norepinephrine-Dopamine Reuptake Inhibitor, SNRI: Serotonin-Norepinephrine Reuptake Inhibitor, SSRI: Selective Serotonin Reuptake Inhibitor, TCA: Tricyclic Antidepressant.

| **Drug** | **Class** | **Subclass** | **Count** |
| --- | --- | --- | --- |
| Antidepressants | | | |
| Sertraline | Antidepressants | SSRI | 81 |
| Escitalopram | Antidepressants | SSRI | 73 |
| Fluoxetine | Antidepressants | SSRI | 29 |
| Vortioxetine | Antidepressants | multimodal | 10 |
| Bupropion | Antidepressants | NDRI | 7 |
| Venlafaxine | Antidepressants | SNRI | 6 |
| Paroxetine | Antidepressants | SSRI | 4 |
| Mirtazapine | Antidepressants | NaSSA | 4 |
| Citalopram | Antidepressants | SSRI | 2 |
| Fluvoxamine | Antidepressants | SSRI | 2 |
| Amitriptyline | Antidepressants | TCA | 2 |
| Mianserin | Antidepressants | NaSSA | 1 |
| Antipsychotics | | | |
| Quetiapine | Antipsychotics | Atypical | 32 |
| Chlorprothixene | Antipsychotics | Typical | 5 |
| Olanzapine | Antipsychotics | Atypical | 4 |
| Aripiprazole | Antipsychotics | Atypical | 2 |
| Flupentixol | Antipsychotics | Typical | 1 |
| Levomepromazine | Antipsychotics | Typical | 1 |
| Mood stabilizers | | | |
| Lamotrigine | Mood stabilizers | Anticonvulsant | 14 |
| Lithium | Mood stabilizers |  | 5 |
| Stimulants | | | |
| Methylphenidate | Stimulants |  | 11 |
| Lisdexamfetamine | Stimulants | Amphetamine | 7 |
| Dexamphetamine | Stimulants | Amphetamine | 1 |
| Anxiolytics | | | |
| Oxazepam | Anxiolytics | Benzodiazepine | 8 |
| Buspirone | Anxiolytics | Azapirone | 1 |
| Pregabalin | Anxiolytics | Gabapentinoid | 1 |
| Alimemazine | Sedatives | Antihistamine | 2 |
| Hydroxyzine | Sedatives | Antihistamine | 2 |
| Melatonin | Sedatives |  | 2 |
| Zopiclone | Sedatives | Z-drugs | 2 |
| Other | | | |
| Sumatriptan | Triptan |  | 1 |

**Table S2: Demographic and Clinical Characteristics: OCD vs Healthy Controls.** "Unknown genetic ancestry" indicates that no specific ancestry group matched perfectly, likely due to mixed genetic ancestry. *Only individuals with OCD whose samples were processed on the same EPIC v2 chips as the healthy controls were included in the comparison. **Genetically predicted ancestry was not available for 248 control individuals; however, their self-reported ancestry was European, and all clustered within the European group in an ancestry PCA. AFR: African, AMR: Native American, CTRL: Healthy Control, EAS: East Asian, EUR: European, OCD: Obsessive-Compulsive Disorder, SAS: South Asian.

|  | OCD (n = 889) | CTRL (n = 384) | Association with case-control status* |
| --- | --- | --- | --- |
| female/male (%female)  *subset for ctrl comparison** | 618/271 (69.5%)  *274/104 (72.5%)* | *275/109 (71.6%)* | *p = 0.672* |
| mean age in years (+/- sd)  *subset for ctrl comparison** | 30.44 (+/- 9.49)  *30.71 (+/- 9.28)* | *30.24 (+/- 6.81)* | *p = 0.293* |
| genetic ancestry | EUR: n = 362  EAS: n = 5  SAS: n = 4  AFR: n = 1  AMR: n = 1  unknown: n = 15  missing: n = 501 | EUR: n = 132  SAS: n = 1  unknown: n = 3  missing: n = 248** | not applicable |
| time points (for saliva collection) | pre-treatment: n = 881  post-treatment: n = 388  follow-up: n = 385 | 1 time point only | not applicable |
| response status  (clinical improvement ≥ 35%) | responder: n = 727  non-responder: n = 162 | not applicable | not applicable |

**Table S3: Baseline Factors Associated with Clinical Response in OCD.** *Directions are described in the main text. GAD: Generalized Anxiety Disorder, OCD: Obsessive-Compulsive Disorder, PTSD: Post-Traumatic Stress Disorder, Y-BOCS: Yale-Brown Obsessive Compulsive Scale.

|  | OCD (n = 889) | Association with response (%improvement)* |
| --- | --- | --- |
| female/male (%female) | 618/271 (69.5%) | **p = 0.030** |
| mean age in years (+/- sd) | 30.44 (+/- 9.49) | **p = 0.001** |
| baseline severity/YBOCS (+/- sd) | 25.82 (+/- 4.19) | p = 0.076 |
| age of diagnosis (+/- sd) | 21.79 (+/- 9.41) | p = 0.448 |
| illness duration in years (+/- sd) | 8.68 (+/- 8.22) | p = 0.535 |
| psychoactive medicine | any: n = 261 (29.4%)  antidepressants: n = 218  antipsychotics: n = 45  mood stabilizers: n = 18  stimulants: n = 18  sedatives: n = 9  anxiolytics: n = 9  missing: n = 63 (7.1%) | any: p = 0.557  antidepressants: p = 0.904  antipsychotics: p = 0.121  mood stabilizers: **p = 0.023**  stimulants: p = 0.130  sedatives: p = 0.619  anxiolytics: p = 0.136 |
| comorbidities | any: n = 332 (37.3%)  GAD: n = 137  depression: n = 115  social phobia: n = 79  panic disorder: n = 68  agoraphobia: n = 45  PTSD: n = 23  anorexia nervosa: n = 11  bulimia nervosa: n = 5  bipolar disorder: n = 5 | any: p = 0.207  GAD: **p = 0.024**  depression: **p = 0.031**  social phobia: p = 0.611  panic disorder: **p = 0.025**  agoraphobia: p = 0.179  PTSD: p = 0.157  anorexia nervosa: p = 0.550  bulimia nervosa: **p = 0.035**  bipolar disorder: p = 0.413 |

**Table S4: Baseline results.** adj: adjusted, agora: agoraphobia, chr: chromosome, DMPs: differentially methylated positions (here top 0.5%), DMR: differentially methylated region, DNAm: DNA methylation, FDR: False Discovery Rate, GAD: generalized anxiety disorder, logFC: log fold change, PTSD: post-traumatic stress disorder, RegulRegion: regulatory region, SE: standard error, SNP: single nucleotide polymorphism, TSS: transcription start site.

**Table S5: Stable results.** adj: adjusted, agora: agoraphobia, chr: chromosome, DMPs: differentially methylated positions (here top 0.5%), DMR: differentially methylated region, DNAm: DNA methylation, FDR: False Discovery Rate, GAD: generalized anxiety disorder, PTSD: post-traumatic stress disorder, RegulRegion: regulatory region, SE: standard error, SNP: single nucleotide polymorphism.

**Table S6: Longitudinal change results.** adj: adjusted, agora: agoraphobia, chr: chromosome, coef: coefficient, DMPs: differentially methylated positions (here top 0.5%), DMR: differentially methylated region, DNAm: DNA methylation, FDR: False Discovery Rate, GAD: generalized anxiety disorder, PTSD: post-traumatic stress disorder, RegulRegion: regulatory region, SE: standard error, TSS: transcription start site.

**Table S7: Cell type proportion results.** agora: agoraphobia, adj: adjusted, GAD: generalized anxiety disorder, PTSD: post-traumatic stress disorder, Resp: Response, SE: standard error.
